# Analysis of the learning curve for Retzius-sparing Robot-assisted radical prostatectomy for a single surgeon

**DOI:** 10.1101/2023.05.16.23290058

**Authors:** H Hussein, N Maitra, J Tay, I Saxionis, R Makin, S Sivathasan, S Smart, A Warren, N Shah, BW Lamb

## Abstract

**Introduction:** The learning curve for retzius sparing robotic radical prostatectomy is not fully understood. This study attempts to identify the learning-curve across the first 130 cases of a single surgeon.

**Methods:** All retzius sparing robotic radical prostatectomy cases performed by a single surgeon between April 2019 and July 2022 were included. Cases were divided chronologically into 3 groups.

**Results:** 130 RS-RARP cases were identified. Statistically significant differences were found between groups in several areas. Median patient age increased between group 1 (59yrs) and Group 3 (66.5yrs) (P=0.04). Proportion of patients with stage >T2 increased between Group 1 (27.9%) and Group 2 (41.9%) (P=0.036). Median console time increased between Group 1 (120 mins) and Group 2 (150 mins,) (P=0.01). Median gland weight increased between Group 1 (28g) and Group 3 (35.5g) (P<0.001). Positive surgical margin rate fell between Group 1 (30.2%) and Group 3 (9.1%).

**Conclusions:** The complexity of cases increased over the learning curve, reflected in older patients, larger prostates and higher stage disease, but the positive surgical margin rate improved with experience. Safety and functional outcomes are excellent throughout. The learning curve might be facilitated by careful case selection favouring smaller prostates with less advanced disease.

## Introduction

Prostate cancer is now considered to be the commonest male cancer in the UK with over 47,000 men diagnosed annually (1). It is considered to be the leading cause of death due to cancer among men in industrialized countries. Prostate-Specific Antigen (PSA) test has become a common tool of screening for prostate cancer in developed countries, increasing the proportion of men diagnosed with potentially curable disease (2). For patients with clinically localized prostate cancer, management options are based on several factors, including risk stratification and life expectancy. Management options for men with potentially curable disease, and life expectancy of a minimum of 10-years include active surveillance, radical prostatectomy, brachytherapy, focal therapies and hormones/external beam radiation therapy (3)

Since its introduction in 2000s, Robotic assisted radical prostatectomy (RARP) has become the standard surgical treatment for clinically localised prostate cancer. However, RARP itself is associated with an increased risk of post-operative treatment-related side-effects, mainly urinary incontinence and erectile dysfunction, which can have a significant impact on the patient’s quality of life, with the highest rates of urinary incontinence usually in the first few months following the radical prostatectomy (4).

With progressive understanding of the regional anatomy of the surrounding structures and the impact of the surgical procedure on them, many trials and surgical techniques had been proposed aiming at maintaining oncological outcomes of the operation while reducing the post-operative sideeffects, mainly urinary incontinence and erectile dysfunction (5).

In 2010, Galfano and his team first reported a new surgical technique (Retzius Sparing Robot Assisted Radical Prostatectomy) aiming to achieve superior post-operative continence rates through preserving the integrity of all the anteriorly located structured such as endopelvic fascia, deep dorsal venous plexus, puboprostatic ligament and the accessory pudendal arteries which are all routinely sacrificed in the conventional anterior approach (6). The superiority of RS-RARP in achieving better functional outcomes earlier has been subsequently validated in several studies compared to the conventional anterior technique (7), however, other studies have found no significant difference in the long-term functional recovery or quality of life outcomes (8). Our own institution has described our early experience of RS-RARP with publication of outcomes compared to a standard RARP approach for the first 22 and 70 cases (9,10).

The early learning curve for RS-RARP has yet to be defined. Galfano’s group found no difference between their first and second 100 RS-RARP patients (6). Analysis of our institutions first 22 and 70 cases found that that outcomes compared favourably with standard RARP from the start (9,10). The aim of the present study was to build on the understanding of the learning-curve for RS-RARP by describing the characteristics and outcomes of the first 130 RS-RARP cases, with specific comparison between the first, second and third groups of this series. Specifically, our objectives were to assess patient characteristics, peri-operative and post-operative patient outcomes across the first 130 cases and to identify if and where learning curves exist.

## Methods

### Patients

All cases of RS-RARP, performed by a single surgeon at a tertiary institution in the period between April 2019 and July 2022 were included. All Retzius-Sparing Robot-Assisted Radical Prostatectomy (RS-RARP) cases were performed during learning curve of a single surgeon who had otherwise performed more than 200 cases of Standard Robot-Assisted Radical Prostatectomy (S-RARP) after completing training in Standard Robot-Assisted Radical Prostatectomy (S-RARP). At the start of the study period, certain RARP procedures performed by this surgeon were by the RS-RARP technique with case selection based on recommended patient specific or disease specific criteria previously described (10, 11). By the end of the study period, all cases performed by the surgeon were RS-RARP.

### Surgical technique and peri-operative management

Our RS-RARP has been previously described and is very similar to that described by Galfano et al. and has previously been described.(6,9) The Da Vinci Si (Intuitive Surgical) was used for all cases. Peri-operative management has also been described and followed BAUS ERAS guidelines (12). First follow up was at 1-week post-op for the removal of catheter, followed by an outpatient follow-up at 6 weeks. PSA levels are done at 6-8 weeks interval from the operation (9,10).

### Data collection

All clinical data was routinely and prospectively entered into patients’ electronic medical record, and subsequently collated by the study team into the study database. Patient demographics included age, body mass index (BMI), PSA, biopsy International Society of Urological Pathology (ISUP) grade group and clinical T-stage (based on MRI). Peri-operative outcomes include type of nerve spare, lymph node dissection, console operative time, estimated blood loss (EBL) and length of stay. Post-operative data included weight of gland, pathological T-stage, ISUP grade group, positive surgical margin (PSM), number of lymph nodes sampled, and post-operative PSA level taken at approximately 6-weeks following surgery. Unplanned admissions and complications within 30-days were recorded. Complications were graded according to Clavien-Dindo system. Erectile function was assessed by patients reported levels as a percentage of pre-operative levels (100% being similar to their pre-operative erectile function status). Continence was assessed by patient reported pad usage numbers.

### Statistical Analysis

All cases were divided chronologically into three groups. Descriptive statistics were calculated for each variable across the total cohort, and for each individual group. Medians and interquartile ranges (IQR) were reported for continuous and categorical variables, respectively. When comparing variables to assess for differences across the three groups, Chi-Squared test was used for categorical data and the Kruskal-Wallis test for continuous data. Where differences were found across the three groups, pairwise comparison was performed between each group to determine where any difference arose. Pairwise comparison was performed using Chi-Squared tests for categorical data and the Mann Whitney U test for continuous data. Statistical significance was taken at the p=0.05 level. Statistical analysis was performed using SPSS 26th Edition, IBM.

## Results

### Patient characteristics

Between April 2019 and July 2022 130 RAR-RARP cases were performed. Cases were divided into three groups, or groups:

1. 43 cases between April 2019 and April 2021
2. 43 cases between April 2021 and December 2021
3. 44 cases between January 2022 and July 2022.

### Pre-operative and intra-operative patient characteristics

Table 1 presents data relating to pre-operative and Intra-operative patient characteristics by operative group. There was no significant difference between groups for BMI, PSA, International Society of Urological Pathology (ISUP) group, nerve sparing status, lymph node dissection status, estimated blood loss, nor intraoperative complications.

**Table 1:**
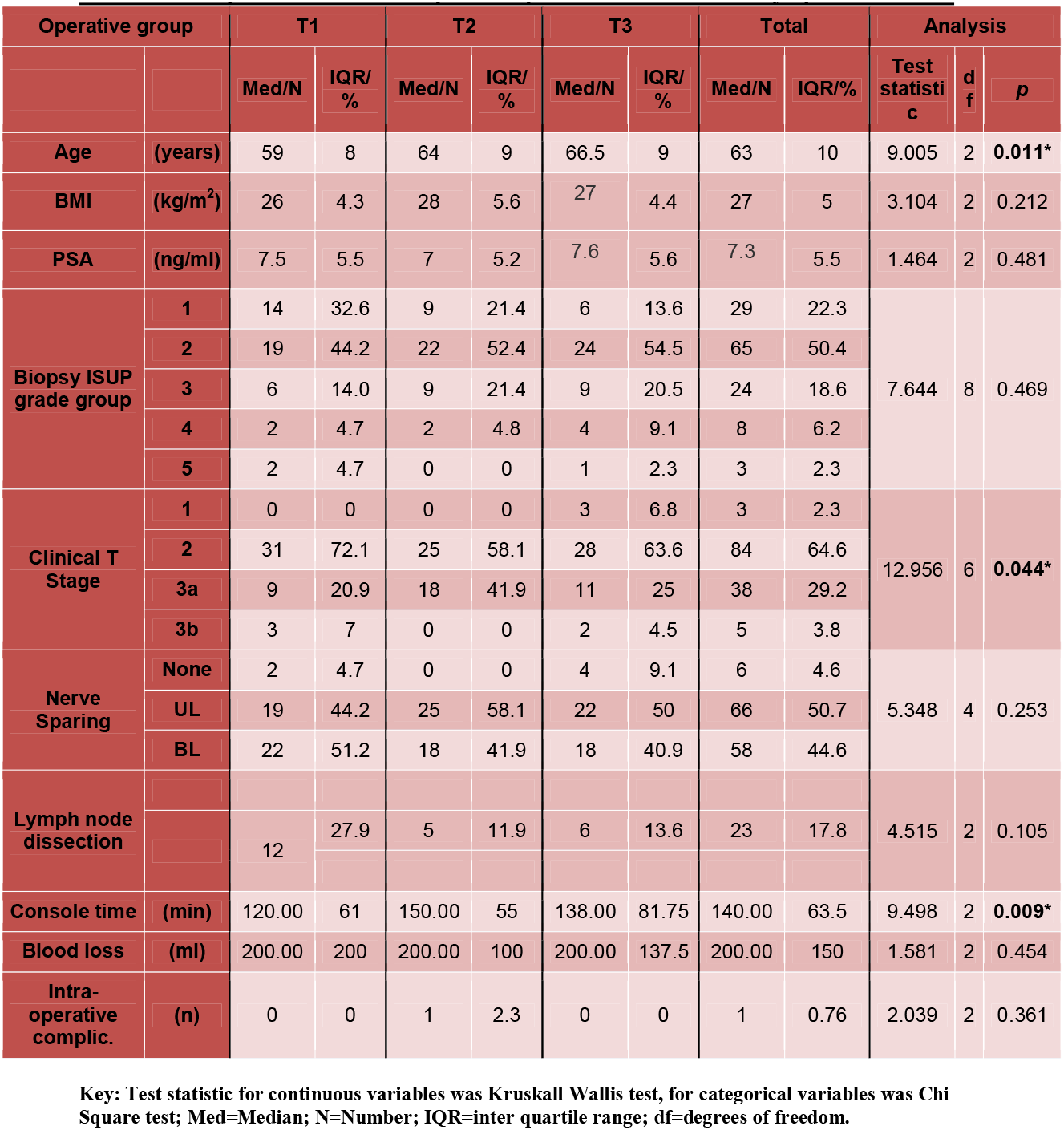
Pre-operative and Intra-operative patient characteristics by operative tertile.

**Table 2:**
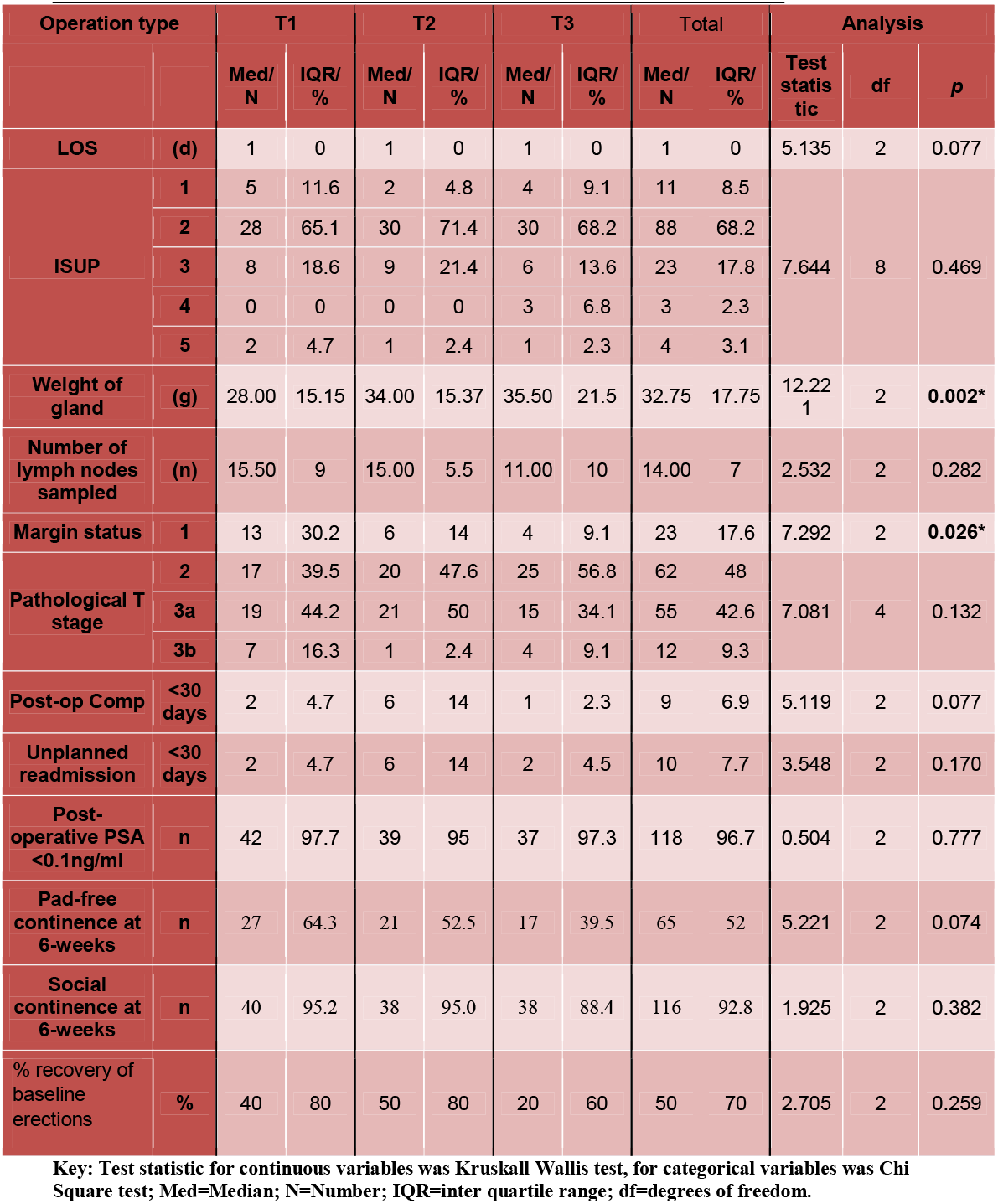
Post-operative characteristics and outcomes by operative tertile.

**Table 3:** Chart displaying results of pairwise comparison for variables with significant differences across tertiles.

|  | Tertiles | Pearson Chi-Square /<br>Mann-Whitney U | Df / Z | P |
| --- | --- | --- | --- | --- |
| **Median Age** | 1 vs. 2 | 727.0 | -1.709 | 0.087 |
|  | 2 vs. 3 | 761.0 | -1.574 | 0.116 |
|  | 1 vs. 3 | 611.0 | -2.848 | **0.004\*** |
| **Proportion of clinical**<br>**T stage >T2** | 1 vs. 2 | 6.643 | 2 | **0.036\*** |
|  | 2 vs. 3 | 6.849 | 3 | 0.077 |
|  | 1 vs. 3 | 3.542 | 3 | 0.315 |
| **Median Weight of**<br>**Gland** | 1 vs. 2 | 666.5 | -1.929 | 0.054 |
|  | 2 vs. 3 | 754.5 | -1.465 | 0.143 |
|  | 1 vs. 3 | 517.5 | -3.513 | **<0.001\*** |
| **Median console time** | 1 vs. 2 | 549.5 | -3.246 | **0.001\*** |
|  | 2 vs. 3 | 838.0 | -0.918 | 0.358 |
|  | 1 vs. 3 | 741.5 | -1.738 | 0.082 |
| **Proportion of**<br>**positive surgical**<br>**margin** | 1 vs. 2 | 3.31 | 1 | 0.069 |
|  | 2 vs. 3 | 0.505 | 1 | 0.477 |
|  | 1 vs. 3 | 6.183 | 1 | **0.013\*** |

A statistically significant difference was found across the groups in the median patient age: T1=59 years, IQR=8; T2=64 years, IQR=9; T3=66.5 years, IQR=9 (KWT=9.005; df=2; p=0.011). Pairwise comparison between groups revealed that a significant difference lay between the median age of groups 1 and 3 (MWU=611.0, Z=-2.848, p=0.004), but not between 1 and 2, or 2 and 3. (Fig. 1)

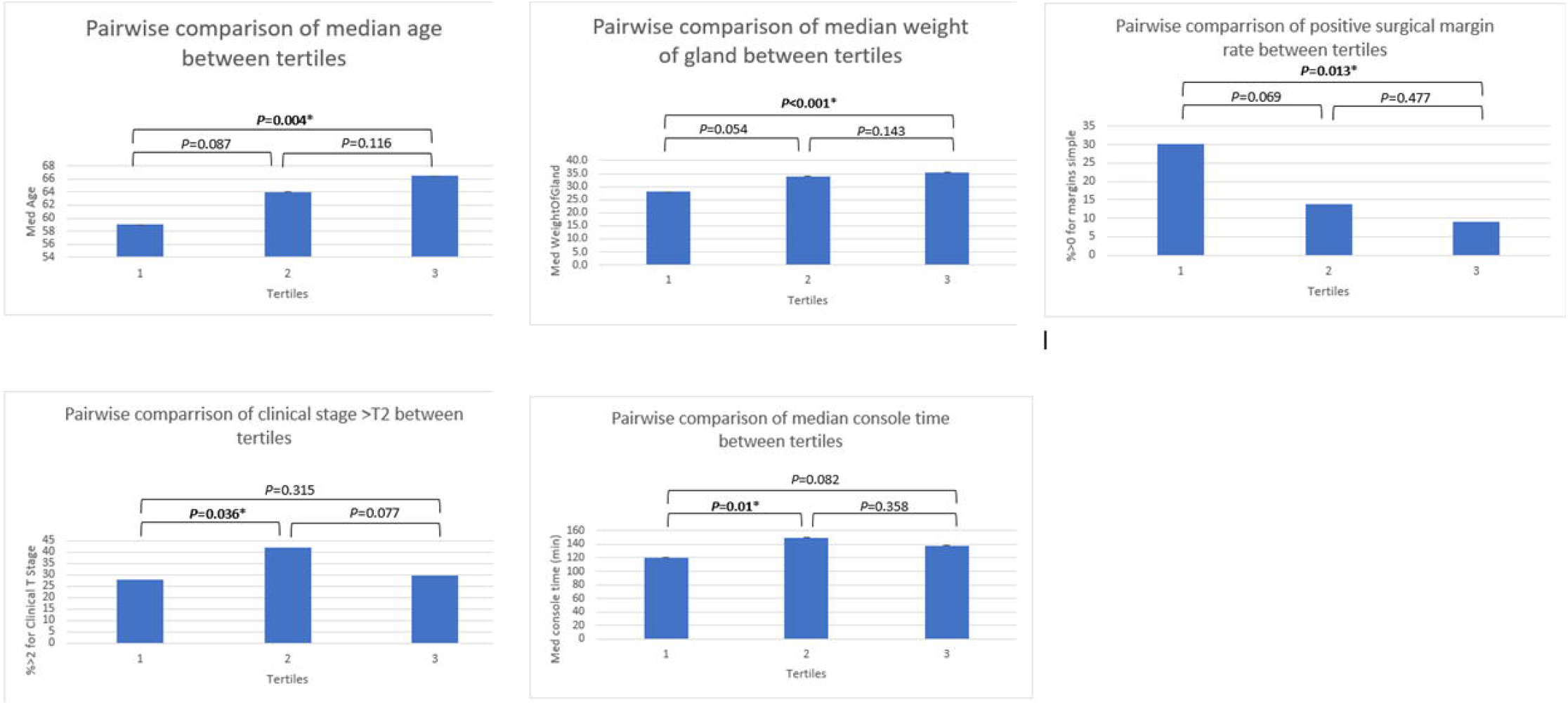

A statistically significant difference was found across the groups in the distribution of clinical T-stage: T1: cT2 n=31, 72%; cT3a=9, 20.9%; T3b n=3, 7%. T2: cT2 n=25, 58.1%; cT3a=18, 41.9%. T3: cT1c=3, 6.8%; cT2 n=28, 63.6%; cT3a=11, 25%; T3b n=2, 4.5%. (CST=12.956; df=6; p=0.044). Pairwise comparison between groups of the proportion of patients with stage >pT2 revealed that a significant difference lay between the median age of groups 1 and 2 (CST=6.643, df=-2, p=0.036), but not between 1 and 3, or 2 and 3. (Fig. 1)

A statistically significant difference was found across the groups in the median console time: T1=120 mins, IQR=61; T2=150 mins, IQR=55; T3=138 mins, IQR=82 (KWT=9.498; df=2; p=0.009). Pairwise comparison between groups revealed that a significant difference lay between the median console time of groups 1 and 2 (MWU=741.5, Z=-1.738, p=0.001), but not between 2 and 3, or 1 and 3. (Fig. 1)

### Post-operative outcomes

Table 1 presents data relating to post-operative outcomes by operative group. There was no significant difference between groups for length of hospital stay, surgical specimen ISUP grade group, number of lymph nodes sampled, pathological T-stage, post-operative complications, unplanned readmissions, post-operative PSA status, pad-free or social continence or recovery of erectile function.

A statistically significant difference was found across the groups in the median post-operative gland weight: T1=28g, IQR=15; T2=34g, IQR=15; T3=35.5g, IQR=21.5 (KWT=12.221; df=2; p=0.002). Pairwise comparison between groups revealed that a significant difference lay between the median console time of groups 1 and 3 (MWU=549.5, Z=-3.513, p=0.001), but not between 1 and 2, or 2 and 3. (Fig. 1)

A statistically significant difference was found across the groups in the positive surgical margin: T1: n=13, 30.2%; T2: n=6, 14%; T3: n=4, 9.1% (CST=7.292; df=2; p=0.026). Pairwise comparison between groups of the proportion of patients with stage >pT2 revealed that a significant difference lay between the median age of groups 1 and 3 (CST=6.183, df=-2, p=0.013), but not between 1 and 2, or 2 and 3. (Fig. 1)

## Discussion

The aim of the current study was to describe the characteristics and outcomes of the first 130 RS-RARP cases performed at our institution; and to draw a comparison between the first, second and third groups of this series. The results demonstrate that patient characteristics were broadly representative of patients undergoing RARP at our institution (9,10). The first group of patients were youngest, with the lowest proportion of clinically locally advanced disease and smallest gland size. The console time was lowest for the first group, and proportion of positive surgical margin highest. The median age and gland size increased progressively over the three groups. The proportion of patient with locally advanced disease increased significantly in the second group, but fell in the third to a level similar to the first. Console time too increased in the second group, then fell in the third, but not all the way to that of the first. The proportion of positive margins fell across the course of the study. There were no other differences in patient characteristics across the study. Complications, length of hospital stay, and unplanned readmissions remained at favourable levels throughout the study. Post-operative PSA levels at 6-weeks were in keeping with the risk profile of operated patients. Functional outcomes were consistent across the study.

As stated previously, there was an intention, as recommended by other authors, early in the RS-RARP learning curve, to select cases with smaller prostate volume, and avoiding anterior tumours (11). It can be seen from the results that the gland size increased across the course of the series, and in the final third (35.5g), was similar to the median gland weight in the previously reported standard RARP cohort (38g) (10). The increase in age seen across the study may reflect the selection, early on in the learning curve, of men with smaller sized prostates, who are likely to be younger (13). Interestingly, console time did not decline across the study, but rose significantly from the first to the second third, before falling in the third. This may also be a reflection of increasing case complexity in the second group, for instance, increased proportion of patients with locally advanced disease and increased gland size whilst maintaining the proportion of patients undergoing lymph node dissection and nerve-sparing. Olivero et al. compared intra- and post-operative outcomes of patients undergoing RS-RARP between experienced and novice RS-RARP surgeons who were on their learning-curve (14). They found that console time was shorter for experienced surgeons, and that novice surgeons performed surgery on patients at an earlier stage. These finding are consistent with those of the present study.

Previously, concerns about the oncological safety of RS-RARP have been raised (8), chiefly because of a higher PSM rate. The PSM rates fell across the course of the present study from 30% in the first third to 9% in the final third. Galfano and colleagues reported an improvement in PSM rates in the second 100 patients (19%) compared to the first 100 patients (32%) in their series (6). Conversely, Olivero and colleagues analysed RS-RARP outcomes performed by surgeons who are on the learning curve, compared to those with >100 cases experience, and concluded that PSM rates and 1 year disease free survival were similar in both groups (14), The proportion of patients in the present study achieving a PSA level of <0.1ng/ml at 6 weeks follow up was high across groups (96.7% overall). These results are important as they support the oncological safety of RS-RARP, in contrast to other studies that have shown an increased in PSM rates, which has led to concerns where surgeons are trying a novel technique. A systematic review by Checcucci and colleagues showed that PSM associated with RS-RARP are often located anteriorly (8). However, careful case selection, avoiding patients with significant anterior index lesions on pre-operative MRI, or other risk factors for locally advanced disease, and those men with smaller prostates may help to avoid an excess risk of PSM early in the learning curve. Our data is consistent with others and suggests that as surgical experience with RS-RARP increases, it might be possible to take on higher risk cases and larger prostates whilst improving oncological outcomes and maintaining excellent functional results (14).

The major benefit for patients of the RS-RARP is the improvement in early continence recovery, which seems consistent across studies. The early experience at our own institution found that continence recovery to be superior to standard RARP within the first 22 cases, with benefits persisting at 12 months (9). The present study found no difference in continence outcomes between groups, with excellent results across the study. The definition of continence is controversial. Although some authors argue that continence should be defined as not requiring any incontinence pads (15), many of our patients report using a security pad despite being continent and describing it a psychologically more reassuring. Accordingly, and in keeping with previously published studies, we included social continence, defined as 0-1 security pad, as a parameter of our outcome as we felt it was clinically more relevant (9,10).

Currently, training for RS-RARP is ad hoc, and to the best of our knowledge, no formalised program of training exists (9). The EAU has developed and implemented a validated surgical curriculum for S-RARP, and training courses and accredited fellowships, such as that at our own institutions, exist (16,17). The lack of a definition of what constitutes best practice may lead to unwarranted variation in quality. RS-RARP experts, along with professional organisations should start by defining best practice, before developing and validating training courses and curriculum for RS-RARP. Such a program might ensure that surgeons are trained to undertake the technique safely and effectively with the shortest possible learning curve.

There are several limitations that should be considered when evaluating the findings of this study. The aim of the current study was to describe the learning curve of RS-RARP. Some outcomes, such as the rate of PSM showed improvement over the 130 included cases, but we do not know if further improvement is likely to be seen, nor if greater numbers of cases were assessed across different surgeons, more subtle elements of the learning curve would have been detected, that perhaps were not apparent in the present analysis.

Secondly, studies have suggested that although there may be an improvement in early continence outcomes in RS-RARP, this benefit is muted by 12 months post-op [18,19]. The outcomes assessed in this study were peri-operative and early post-operative outcomes. Further work is needed to assess the learning curve on longer term outcomes, such as BCR, sexual function, urinary continence and quality of life.

Thirdly, self-reported erectile function was not measured with a validated questionnaire for all patients. Due to the covid-19 pandemic many follow-up consultations were undertaken virtually, which made it difficult to complete comprehensive patient reported outcome measures.

Finally, as a single surgeon series, the findings in the current study may not be entirely generalisable. Further research is needed to assess the learning curves and longer-term oncological and functional outcomes, across a number of surgeons and institutions, before the findings can be confidently generalised.

## Conclusion

The present study suggests that RS-RARP has a learning curve, which could be augmented by careful case selection, starting with smaller volume prostates and avoiding significant anterior lesions. We found that that the rate of positive surgical margins improves with experience, but that safety and functional outcomes are excellent from early in the learning curve. Further work is needed to develop formalised training curricula and a structured training for surgeons wishing to learn this novel and potentially advantageous technique.

## Data Availability

All data produced in the present study are available upon reasonable request to the authors

## Acknowledgements

Urology Department Addenbrooke’s Hospital, Cambridge University Hospitals

## Notes

Conflicts of interest: all authors confirm they have no conflict of interest that are relevant to the content of this article

### Competing Interest Statement

all authors confirm they have no conflict of interest that are relevant to the content of this article

### Funding Statement

This study did not receive any funding

### Author Declarations

Ethics committee/IRB of Cambridge University Hospitals NHS Foundation Trust waived ethical approval for this work

